# Characteristics and Attitudes of Wearable Device Users and Non-Users in a Large Healthcare System

**DOI:** 10.1101/2023.08.10.23293960

**Authors:** Rachael A. Venn, Shaan Khurshid, Mia Grayson, Jeffrey M. Ashburner, Mostafa A. Al-Alusi, Yuchiao Chang, Andrea Foulkes, Patrick T. Ellinor, David D. McManus, Daniel E. Singer, Steven J. Atlas, Steven A. Lubitz

## Abstract

**Introduction:** Consumer wearable devices with health and wellness features are increasingly common and may enhance prevention and management of cardiovascular disease. However, the characteristics and attitudes of wearable device users versus non-users are poorly understood.

**Methods:** Wearable Activity Tracking for Comprehensive Healthcare-Integrated Technology (WATCH-IT) was a prospective study of adults aged ≥18 years receiving longitudinal primary or ambulatory cardiovascular care at one of eleven hospitals within the Mass General Brigham multi-institutional healthcare system between January 2010-July 2021. We invited patients, including wearable users and non-users, to participate via an electronic patient portal. Participants were asked to complete a 20-question survey regarding perceptions and use of consumer wearable devices. Responses were linked to electronic health record data. Multivariable logistic regression was used to identify factors associated with device use.

**Results:** Among 280,834 individuals receiving longitudinal primary or cardiovascular care, 65,842 did not have an active electronic portal or opted out of research contact. Of the 214,992 individuals sent a survey link, 11,121 responded (5.2%), comprising the WATCH-IT patient sample. Most respondents (55.8%) reported current use of a wearable device, and most non-users (95.3%) reported they would use a wearable device if provided at no cost. Although most users (70.2%) had not shared device data with their doctor previously, the majority believed it would be very (20.4%) or moderately (34.4%) important to share device-related health information with providers. In multivariable models, older age (odds ratio [OR] 0.80 per 10-year increase, 95% CI 0.77-0.82), male sex (0.87, 95% CI 0.80-0.95), and heart failure (0.75, 95% CI 0.63-0.89) were associated with lower odds of wearable device use, whereas higher median zip code income (1.08 per 1-quartile increase, 95% CI 1.04-1.12) and care in a cardiovascular medicine clinic (1.17, 95% CI 1.05-1.30) were associated with greater odds of device use. Nearly all respondents (98%) stated they would share device data with researchers studying health outcomes.

**Conclusions:** Within an electronically assembled cohort of patients in primary and cardiovascular medicine clinics with linkage to detailed health records, wearable device use is common. Most users perceive value in wearable data. Our platform may enable future study of the relationships between wearable technology and resource utilization, clinical outcomes, and health disparities.

---

Consumer wearable technologies are increasingly common, with nearly one third of Americans reporting use of a smart watch or fitness tracker. ^1,2^ Wearable devices are commonly marketed to promote general health and wellness, and many commercially available products measure physiologic parameters with established clinical and prognostic significance (e.g., heart rate,^3^ physical activity levels^4,5^). Selected wearable devices even contain the ability to record data previously limited to the domain of medical diagnostics, such as single-lead electrocardiograms.

Importantly, data regarding real-world use of wearable devices in the healthcare setting remains limited. Results from small prospective studies^6,7^ and a recent survey^2^ suggest that patterns of use may differ by patient comorbidities and that data collected from wearable devices may influence patient-physician interactions.^7^ Understanding the characteristics of wearable device users and their attitudes toward medical applications of device use will be foundational in ensuring equitable access to wearable technology, identifying specific barriers to wearable use, and clarifying the potential role of wearables for healthcare delivery. To this end, creation of a sizeable cohort of wearable device users within a large healthcare system, with the potential to link device data with clinical information within the electronic health record (EHR), may serve as a unique resource to evaluate how wearable device data can be used to improve clinical outcomes.

The Wearable Activity Tracking for Comprehensive Healthcare-Integrated Technology (WATCH-IT) study is a prospective study of ambulatory patients within a large multi-institutional healthcare system. Here, we report the clinical characteristics of the WATCH-IT cohort, including factors associated with wearable device use, and the results of a survey assessing patient attitudes towards health-related applications of wearable devices.

## Methods

### Study design and patient cohort

In WATCH-IT, we electronically assembled a cohort of patients to enable study of wearable device users, including wearable device data and health outcomes. We derived the candidate list of study participants from a) the Community Care Cohort Project (C3PO),^8^ a previously described cohort comprising over 500,000 adults aged ≥18 years receiving longitudinal primary care at one of eleven hospitals within the Mass General Brigham (MGB) network, and b) the Enterprise Warehouse of Cardiology (EWOC), an analogous cohort comprising individuals receiving longitudinal cardiovascular care.^9^ Individuals included in both C3PO and EWOC have clinical EHR data available within a centralized warehouse. To select for patients most likely to be active within the MGB system, both cohorts were filtered to identify patients who were alive and had start of clinical follow up (i.e., the date of the second primary care visit of the earliest qualifying pair) after 1/1/2010.^8^ The resulting list of roughly 300,000 patients was uploaded into a custom tool developed for Epic (Verona, WI), the clinical EHR platform utilized by MGB. Patients were matched to the complete MGB Epic database using dual identifiers including medical record number and date of birth. Once uploaded into the EHR template, active users of the MGB electronic patient portal were identified, facilitating direct-to-patient messaging via the Epic platform.

After identifying registered users of the electronic patient portal and excluding individuals who had previously opted out of research communications, we used the portal messaging system to send electronic invitations to participate in the WATCH-IT survey (date range of invitations 11/2021-4/2022). The initial invitation letter included a direct link to the WATCH-IT survey instrument (**Supplemental Figure 1**). Survey non-responders were sent a reminder letter up to two weeks after initial contact date. Upon completion of the survey, participants were entered into a lottery for an Amazon gift card in the amount of $25. Survey respondents provided informed consent and all study protocols were approved by the MGB Institutional Review Board.

### Survey instrument

The survey instrument was built and managed using REDCap (Research Electronic Data Capture), a secure, web-based software platform designed to support data capture for research studies.^10,11^ The survey utilized branching logic with initial bifurcation based on active use of a wearable device, defined as a “Yes” response to the survey question “Do you currently use a wearable device?” (**Figure 1** and **Supplemental Figure 1**). Individuals who opened a survey but not answer this initial question were classified as non-responders. Users were then asked a series of questions regarding patterns of current device use, while non-users (defined as a “No” response to the question above) were asked about access to wearable device technology and reasons for non-use. We defined a subcategory of non-users, called “potential users,” who reported they would use a wearable device if one were provided at no cost (**Figure 1**). Users and potential users were asked about potential medical applications of device use, and all respondents were asked a series of general health-related questions.

**Figure 1.**
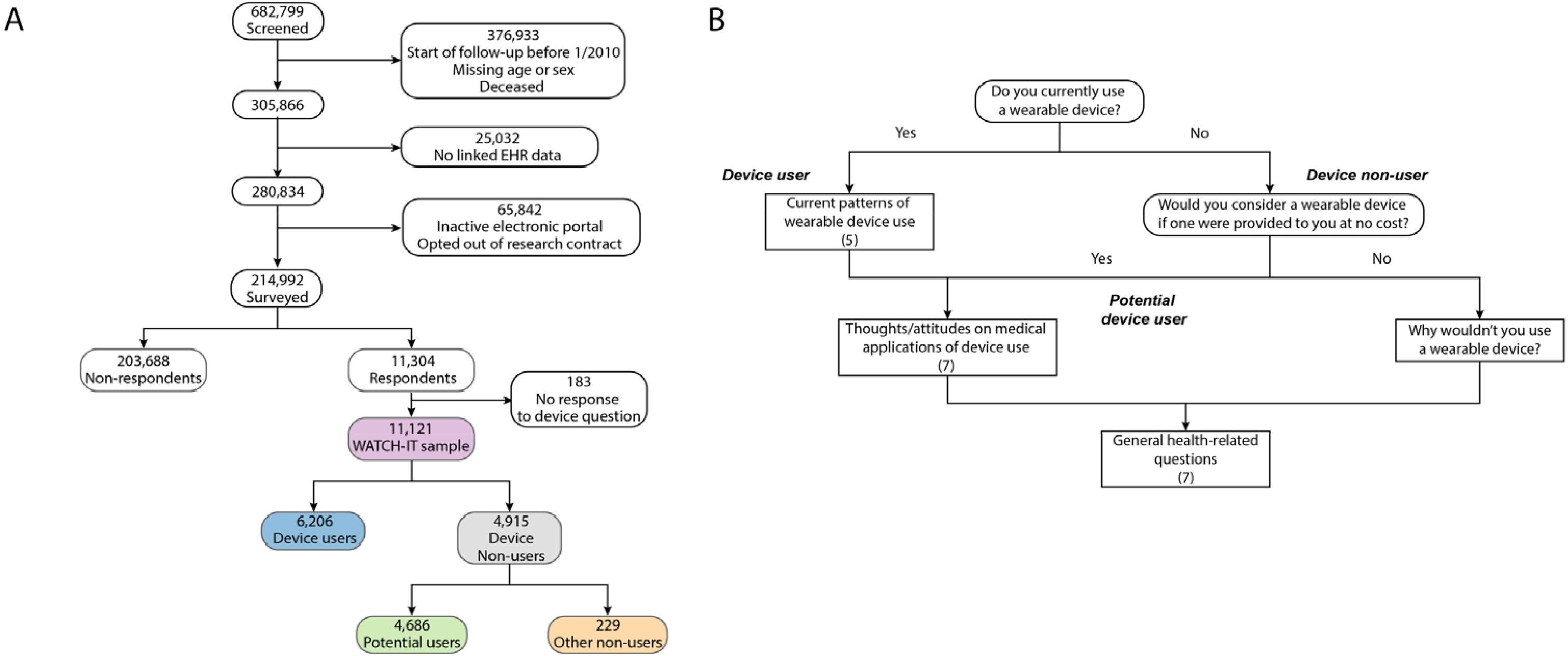
Study summary. Depicted is an overview of sample construction (panel A) and the survey instrument (panel B). Using the electronic health record (EHR), we screened a total of 682,799 individuals with at least two visits within 1-3 years in a cardiology or primary care practice. Of these, a total of 214,992 were contacted and received a survey instrument. Of the individuals contacted, 11,121 responded to at least the initial question, comprising the WATCH-IT sample. Of these, individuals were then classified as device users versus non-users, as described in panel B. Samples comprising the main analyses sets are highlighted in color. The short WATCH-IT survey comprised branching questions (rounded rectangles) and non-branching questions (rectangles). Responses to branching questions served to classify individuals as device users, device non-users, and potential device users, and impacted which questions were administered.

Survey respondents self-identified by name and date of birth, which were used to link responses to complete health information contained in the EHR. Surveys which could not be linked by this mechanism were excluded from analysis (**Figure 1**).

### Clinical factor ascertainment

Demographic and clinical characteristics were derived from the EHR. Demographic data included age, sex, race, and ethnicity, and was extracted from demographic fields within the patient chart. Race was classified as White, Black, Asian, other race, or unknown race in accordance with available fields within the EHR. Clinic membership (primary care versus cardiology) was assigned according to whether individuals were first identified from the primary care versus cardiovascular parent samples. Median income was assigned based on ZIP code according to 5-year US Census data from 2017-2021 (<u>data.census.gov</u>, accessed 11/02/2022), and then divided into quartiles. For 131 individuals (1.1%) without income data, the sample median income was assumed. Patient portal contact was quantified as the number of unique interactions with patient messages or results within the preceding year, which was then categorized into approximate quartiles (i.e., 0, 1-4, 5-19, ≥20). Prevalent diseases were defined based on composite data comprising International Classification of Diseases, 9^th^ and 10^th^ revision (ICD-9 and 10) diagnosis codes, Current Procedural Terminology (CPT) codes, and medications. Codes used to define atrial fibrillation, coronary disease, diabetes, hyperlipidemia, hypertension, heart failure, peripheral arterial disease, and stroke have been previously validated (positive predictive value ≥85%).^8,12,13^ All disease definitions are provided in **Supplementary Table 1**.

### Statistical analysis

The distribution of survey responses was plotted using bar graphs. Question-specific non-response rates ≥0.5% were plotted as separate bars. Associations between clinical factors and wearable device use were assessed using multivariable logistic regression, with device use as the outcome and age, sex, race, income, clinic membership (primary care versus cardiovascular medicine), portal contact, and presence of atrial fibrillation, coronary disease, diabetes, hyperlipidemia, hypertension, heart failure, other arrhythmias, obesity, peripheral artery disease, pulmonary disease, and stroke as covariates. Clinical covariates were selected *a priori* based on hypothesized potential associations with device use. Ethnicity was not included in the models given a low frequency of Hispanic individuals (<1%). We then fit analogous models with device use (versus potential device use), potential device use (versus no potential device use), and survey response (versus non-response) as alternative outcomes including the same adjustment variables.

We considered two-sided p-values <0.05 to indicate statistical significance. All analyses were performed using R v4.0 (packages ‘data.table’, ‘plyr’, and ‘stringr’).

## Results

A total of 280,834 individuals met criteria for longitudinal primary or ambulatory cardiovascular care. Of these, 65,842 patients were excluded for either inactive electronic portal or having opted out of research contact, resulting in 214,992 individuals who were sent a survey invitation. Of invited patients, 11,121 at least partially completed a survey (5.2% response rate). Survey respondents comprised the WATCH-IT sample, with mean age 57.6 years (standard deviation 15.4), 62.9% women, and 92.5% White. Complete baseline characteristics of the WATCH-IT cohort are provided in **Table 1**. Characteristics of survey responders versus non-responders are provided in **Supplementary Table 2**.

**Table 1.** Baseline characteristics of the primary analysis sample.

| Characteristic | Survey respondents |  |  |
| --- | --- | --- | --- |
|  | Device users<br>(n=6,206) | Device non-users<br>(n=4,915) | Overall<br>(n=11,121) |
| Age, years | 55.3 ± 15.5 | 60.4 ± 14.8 | 57.6 ± 15.4 |
| Female, n (%) | 4,078 (65.7%) | 2,918 (59.4%) | 6,996 (62.9%) |
| Race, n (%)<sup>*</sup> |  |  |  |
| Asian | 181 (2.9%) | 155 (3.1%) | 336 (3.0%) |
| Black | 193 (2.9%) | 149 (3.0%) | 342 (3.0%) |
| Other | 78 (1.3%) | 45 (0.9%) | 123 (1.1%) |
| Unknown | 17 (0.3%) | 20 (0.4%) | 37 (0.3%) |
| White | 5,737 (92.4%) | 4,546 (92.5%) | 10,283 (92.5%) |
| Ethnicity, n (%) |  |  |  |
| Hispanic | 53 (0.9%) | 29 (0.6%) | 82 (0.7%) |
| Non-Hispanic | 5,395 (86.9%) | 4,306 (87.6%) | 9,701 (87.2%) |
| Unknown | 758 (12.2%) | 758 (15.4%) | 1,338 (12.0%) |
| Median ZIP code income, \$ (quartile 1, quartile 3) | 104,209<br>(84,354, 127,646) | 102,088<br>(81,027, 124,528) | 102,672<br>(83,469, 126,185) |
| Cardiology clinic, n (%) | 2,242 (36.1%) | 1,962 (39.9%) | 4,204 (37.8%) |
| Median patient portal contacts (quartile 1, quartile 3)<sup>†</sup> | 4 (0, 18) | 5 (0, 19) | 4 (0, 18) |
| Obesity, n (%) | 1,816 (29.3%) | 1,472 (29.9%) | 3,288 (29.6%) |
| Hypertension, n (%) | 2,973 (47.9%) | 2,845 (57.9%) | 5,818 (52.3%) |
| Diabetes, n (%) | 1,068 (17.2%) | 1,045 (21.3%) | 2,113 (19.0%) |
| Hyperlipidemia, n (%) | 3,391 (54.6%) | 3,165 (64.4%) | 6,556 (59.0%) |
| Coronary disease, n (%) | 1,624 (26.2%) | 1,595 (32.5%) | 3,219 (28.9%) |
| Heart failure, n (%) | 320 (0.5%) | 380 (0.8%) | 700 (6.3%) |
| Pulmonary disease, n (%) | 1,998 (32.2%) | 1,686 (34.3%) | 3,684 (33.4%) |
| Atrial fibrillation, n (%) | 1,003 (16.2%) | 883 (18.0%) | 1,886 (17.0%) |
| Other arrhythmia, n (%) | 1,941 (31.3%) | 1,681 (34.2%) | 3,622 (32.6%) |
| Stroke, n (%) | 519 (8.4%) | 549 (11.2%) | 1,068 (9.6%) |
| *Reflects self-reported race. Category Other includes American Indian or Alaskan Native, Native Hawaiian or other Pacific Islander, or individuals reporting two or more race categories, and individuals reporting a race not included among the listed categories. |  |  |  |
| †Number of interactions with results or messages using patient portal in the year preceding WATCH-IT study period |  |  |  |

Of the 11,121 individuals in the WATCH-IT sample, most (83.1%) reported prior use of telehealth visits and/or use of an online portal for communication with healthcare providers. More than half of survey respondents (n=6,206, 55.8%) reported current use of a wearable device. Among device non-users (n=4,915), almost all (n=4,686, 95.3%) would use a wearable device if one were provided at no cost (classified as potential users). Only 7.8% of all respondents reported that a doctor had previously recommended use of a wearable device for medical reasons (**Supplemental Figures 2-3**).

Of the 6,206 device users, most (72.0%) stated they wore their device for at least 12 hours per day in the preceding seven days. Most reported use of either an Apple (56.2%) and Fitbit (28.2%) brand device (**Supplemental Figure 4**). Whereas only 24.8% of device users had previously shared device data with their doctors, more than half (54.8%) felt it was important for healthcare providers to be aware of information collected by their device (**Supplemental Figure 5**). Users identified the following categories as particularly important to share with providers: heart rate (selected by 89.7% of users), activity levels (64.4%), health conditions (53.6%), pulse oximetry (60.0%), and sleep (49.3%) (**Supplemental Figure 5)**. Most users agreed (93.7%) that they would get peace of mind knowing their device could detect a heart problem if they had one, and only a small minority (8.8%) reported anxiety related to potential detection of unknown conditions (**Supplemental Figure 6**). After receipt of an abnormal notification from their device, 87.7% of users would contact their medical provider as a next step (**Supplemental Figure 7**). Almost all users (91.2%) would be willing to share information from their device with researchers studying health outcomes (**Supplemental Figure 8**). Responses were generally consistent for potential users (**Supplemental Figures 10-12**).

In multivariable models evaluating factors predictive of wearable device use (versus non-use), greater median zip code income (1.08 per 1-quartile increase, 95% CI 1.05-1.12), history of atrial fibrillation, and receipt of cardiovascular care (1.16, 95% CI 1.04-1.29) were independently associated with greater odds of device use; whereas older age (odds ratio [OR] 0.80 per 10-year increase, 95% CI 0.77-0.82), male sex (OR 0.86, 95% CI 0.79-0.93), and heart failure (0.75, 95% CI 0.63-0.89) were associated with lower odds of device use (**Figure 3**). Similar associations were observed when comparing device users to potential users (**Figure 3**). Among non-users, older age, male sex, and White race were associated with lower odds of being a potential consumer wearable device user, whereas obesity, hyperlipidemia, pulmonary disease and cardiology patient status were associated with greater odds of being a potential user (**Figure 3**). When assessing factors associated with WATCH-IT survey response, older age, male sex, higher income, hypertension, heart failure, peripheral arterial disease, and stroke were associated with lower odds of survey response, whereas White race, cardiology patient status, greater patient portal contact, atrial fibrillation, hyperlipidemia, other arrhythmias, and obesity were associated with greater odds of survey response (**Figure 3**).

**Figure 2.**
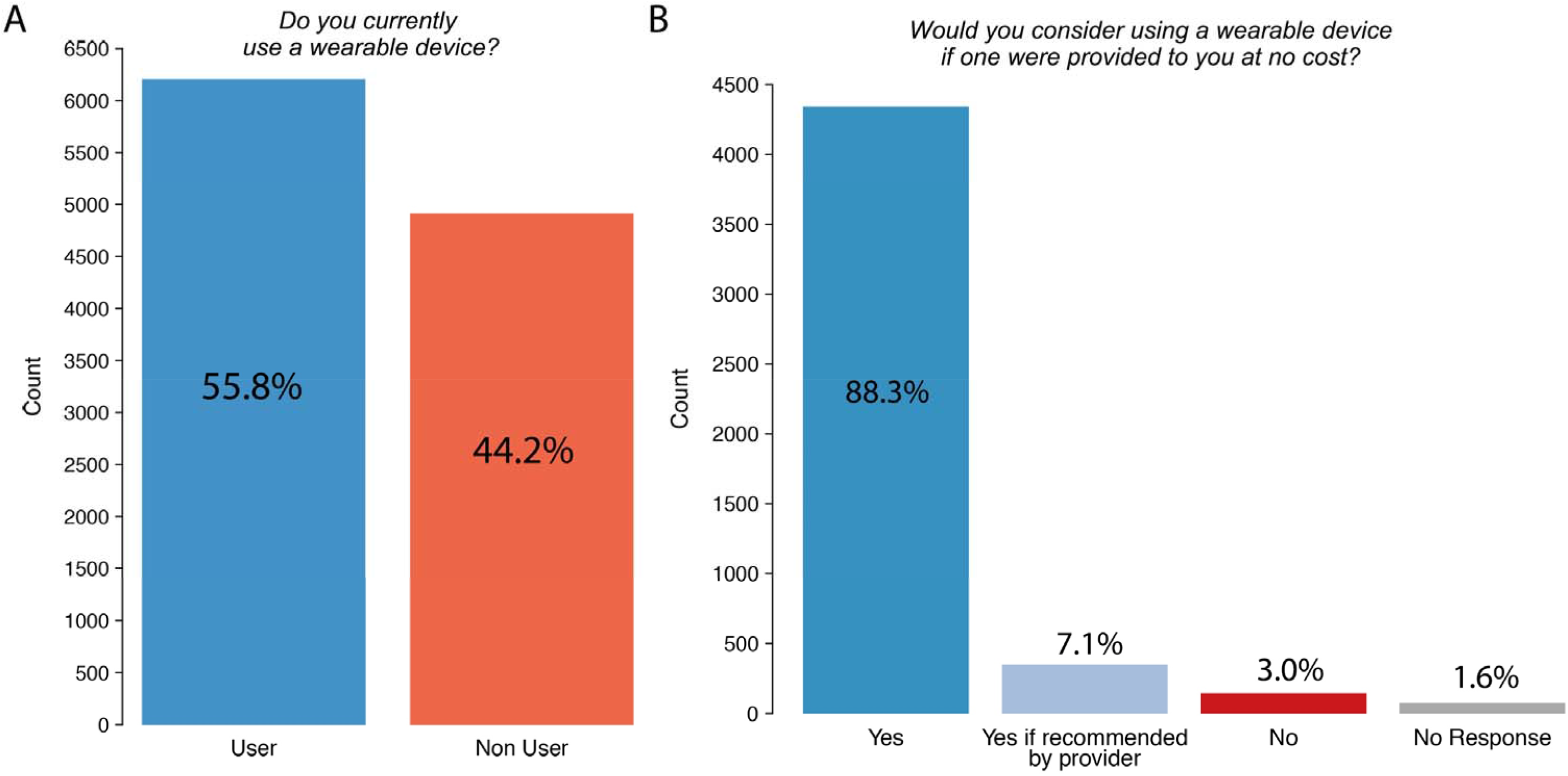
Frequency of active and potential wearable device use. Depicted is the frequency of actual and potential wearable use in the analysis sample. Panel A depicts the distribution of responses to the question “Do you currently use a wearable device?” Panel B depicts the distribution of responses to the question “Would you consider using a wearable device if one were provided to you at no cost?”

**Figure 3.**
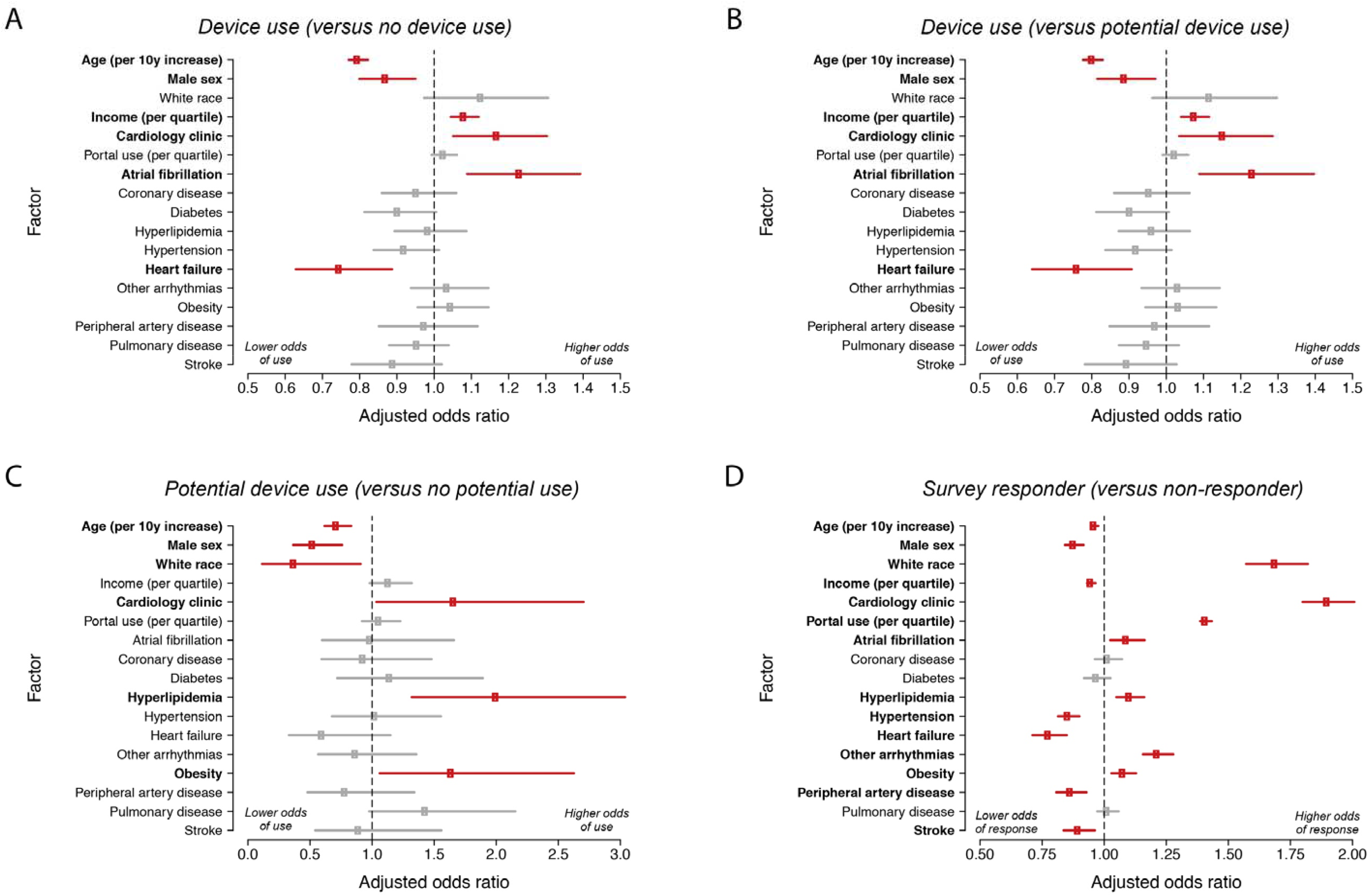
Factors associated with device use, potential device use, and survey response. Plotted are results of multivariable logistic regression with the following outcomes: wearable device use versus non-use (panel A), wearable device use versus potential device use (panel B), potential device use versus no potential device use (panel C), and survey response versus non-response (panel D). Each model is adjusted for each of the listed factors on the y-axis as covariates. Factors with significant associations are bolded and plotted with red points (versus gray points). Error bars indicate 95% confidence intervals.

## Discussion

In the current study, we leveraged direct-to-patient communication at scale to develop a novel cohort of over 11,000 primary care and cardiovascular patients across a large multi-institutional healthcare system with linked EHR data. We found that roughly half of participants reported active use of a consumer wearable device. Atrial fibrillation and cardiology patient status were associated with greater odds of wearable device use; while older age, male sex, lower income, and heart failure were associated with lower odds of wearable device use.

Our study supports and extends prior work characterizing patterns and attitudes related to health technology and wearable device use. A recent analysis of approximately 2,000 individuals from the Framingham research cohort found that age, sex, and educational attainment were each associated with digital health engagement.^6^ A second survey-based study including just over 1,200 respondents reported that patients with atrial fibrillation were more likely to share wearable device data with their healthcare providers.^14^ A nationally representative survey relying solely on self-reported data found that an estimated 29% of individuals use wearable devices, and that those with cardiovascular conditions had a lower prevalence of device use.^2^ In our comparably large sample of individuals with linked electronic health record data, we found that device use is common, and that demographic and clinical factors such as age, sex, income, clinic membership, and cardiac comorbidities such as atrial fibrillation and heart failure substantially impacted the odds of wearable device use.

Our results have several implications. First, our findings suggest a discordance between the high health-related value of wearable technology perceived by patients versus their relatively limited use and uncertain clinical utility in current clinical practice. Specifically, survey respondents felt that device-based monitoring of physiologic parameters such as heart rate and pulse oximetry offered peace of mind and provided important information to share with healthcare providers. Despite this, only a minority of wearable device users had previously shared device data with their doctors. Additionally, while very few survey respondents reported that their doctors had recommended wearable device monitoring, most device users would contact their healthcare provider as a next step after receiving an abnormal device alert.

Potential barriers to the implementation of wearable devices as health tools may include absence of rigorous evidence of therapeutic benefit, patient or provider unfamiliarity with specific devices and their capabilities, and a relative lack of consensus guidelines defining appropriate use of wearable devices in a clinical setting.^15^ Of note, while the ability to collect a wide range of biophysical data for longer durations and across broader patient populations offers a potential advantage of consumer-based wearable devices over traditional forms of ambulatory monitoring (e.g., patch monitors, blood pressure recording devices, daily weights), provider bandwidth and lack of accepted reimbursement models for interpreting such data may serve as key barriers. There are also important uncertainties regarding potential provider liability for acting on wearable-based data and a general absence of ready infrastructure for integrating wearable-based data into existing electronic health record platforms and clinical workflows.^16,17^ To this end, future prospective studies powered to demonstrate clinical benefit and evaluate strategies for integrating wearable device data into clinical practice will be critical to ensure that the patient-perceived value of wearable devices is supported by robust evidence and commensurate with their true utility.

Second, our analyses suggest that careful attention to specific demographic, socioeconomic, and clinical factors influencing wearable device use will be critical in ensuring equitable access and distribution of care. The potential for clinical use of wearable devices to worsen existing health disparities is supported by our finding that over 95% of survey respondents identified as non-users reported that they would use a wearable device if it were offered at no cost. Additionally, in models adjusted for clinical variables, the odds of device use were substantially higher among individuals living in zip codes with greater income, while patient-specific factors previously associated with disparate access to care and poor health outcomes, including older age and the presence of heart failure, were associated with lower odds of device use. Our models suggest that similar factors were associated survey response itself, suggesting the importance of sociodemographic factors and comorbidities may be even greater in the general population. Additional work is warranted to better understand potential barriers to access in vulnerable patient populations and to ensure that future initiatives leveraging mobile technology to improve health outcomes are implemented equitably.

Third, we submit that the WATCH-IT study provides an important proof-of-concept for electronic contact deployed at scale to assemble a cohort of wearable device users with linked EHR data for outcomes research. Large-scale assessments of wearable technology, such as the Apple Heart Study^18^ and Fitbit Heart Study,^19^ each using photoplethysmography (PPG) to detect heart rate and rhythm through variation in pulse interval, have been limited by an inability to link wearable device data to detailed health information. Conversely, smaller assessments in well-curated research cohorts^6^ are limited by modest sample sizes and are subject to healthy volunteer bias. WATCH-IT complements prior approaches by providing a large sample of device users engaged with longitudinal ambulatory care, through which robust clinical EHR data are regularly ascertained. To this end, we importantly observed that nearly all respondents reported willingness to share device-based information with researchers studying health outcomes. Samples such as the WATCH-IT cohort may provide a ready platform for rigorous assessment of future wearable device-based interventions with sufficient power to detect associations with longitudinal outcomes.

Our study should be interpreted in the context of design. First, our observations are limited to portal users and survey respondents. We submit that bias is inherent in survey-based approaches and a 5% response rate is typical for electronically-delivered surveys.^20^ Furthermore, availability of linked EHR data in the parent samples comprising WATCH-IT allowed us to define specific factors associated with survey response (e.g., younger age, White race, cariology clinic membership), allowing us to better understand the potential effects of response bias in our study. Second, although we used previously validated algorithms to define clinical factors, some degree of misclassification of exposures remains likely. Third, most individuals included were White, and there was minimal representation of Hispanic individuals. While the absolute number of individuals in our study from racial minority groups compares favorably to prior studies of wearable technology,^6,14^ our findings may not generalize to populations with varying racial and ethnic composition.

In summary, the WATCH-IT study utilized a scalable platform of EHR-based communication to prospectively enroll over 11,000 patients receiving longitudinal primary or cardiovascular care, finding that use of consumer-based wearable devices is common within this cohort across a large healthcare system. Interest in clinical usage of wearable devices was nearly universal, and attitudes toward sharing device-related data were positive. Future research leveraging samples like the WATCH-IT cohort could empower rigorous investigation of associations between wearable device use and longitudinal clinical outcomes and provide a platform to evaluate mobile-health based interventions.

## Data Availability

Raw identifiable data included in this manuscript are not available due to privacy considerations.

